# Diagnostic performance of urinary 5-Hydroxyindoleacetic Acid in acute appendicitis: a systematic review and diagnostic test accuracy meta-analysis

**DOI:** 10.1101/2023.08.02.23293546

**Authors:** Javier Arredondo Montero, Oscar Emilio Bueso Asfura, Blanca Paola Pérez Riveros, María Rico Jiménez

## Abstract

**Background:** This study aimed to analyze the diagnostic performance of urinary 5-Hydroxyindoleacetic Acid (5-HIAA) in acute appendicitis (AA).

**Methods:** We conducted a systematic review of the literature in the main databases of medical bibliography. Two independent reviewers selected the articles and extracted relevant data. Methodological quality was assessed using the QUADAS2 index. A synthesis of the results, standardization of the metrics, and a random-effect meta-analysis were performed. Additionally, a diagnostic test accuracy meta-analysis (DTA) was performed.

**Results:** Twelve studies with data from 1467 participants (724 patients with a confirmed diagnosis of AA and 743 controls) were included in this review. The random-effect meta-analysis of urinary 5-HIAA (AA vs controls) included 7 articles (352 AA and 258 controls) and resulted in a significant mean difference [95% CI] of 23.30 [15.82-30.77] μmol/L (p<0.001). The DTA meta-analysis of urinary 5-HIAA included 8 articles and resulted in a pooled sensitivity [95% CI] of 68.6 [44.1-85.9]% and a pooled specificity [95% CI] of 82 [54.7-94.5]%.

**Conclusions:** Although the evidence is heterogeneous and limited, urinary 5-HIAA emerges as a potential non-invasive diagnostic tool for AA. Urinary 5-HIAA does not seem to be a useful biomarker to distinguish between NCAA and CAA. Future prospective studies with a large sample size and a rigorous design are necessary to validate these findings.

## Introduction

Acute appendicitis (AA) is the most frequent cause of acute surgical abdomen worldwide [1]. At present, AA continues to be a major problem today in terms of social and health care, both because of the high rate of diagnostic error and because of the human and economic costs derived from the latter [1–3]. Many efforts have been made in the last decades to optimize the diagnostic performance of AA. This has led to the appearance of promising biomarkers, such as Interleukin-6 [4] and novel blood count ratios (i.e platelet-to-lymphocyte ratio, or systemic-immune inflammation index) [5,6] that improve parameters classically used in isolation, such as leukocytosis and neutrophilia. The same has happened with the appearance of new multivariate indexes that have improved the diagnostic performance of the classic Alvarado score [7,8]. The creation of standardized programs for reporting ultrasound evaluations has improved the already excellent diagnostic performance of this tool [9,10]. However, ultrasound is an operator-dependent tool and therefore cannot be considered a unique and perfect diagnostic test at present. Finally, it is pertinent to highlight the promising role of artificial intelligence and predictive models in this pathology, with growing evidence in recent times [11].

Regarding the non-invasive diagnosis of AA, the existing literature is scarce. Recently, a systematic review and meta-analysis on the role of urinary LRG1 as a diagnostic test in pediatric AA was published, and although the evidence was heterogeneous and limited, it appears that this molecule is emerging as a promising diagnostic test in this subpopulation [12].

The cecal appendix is enriched with enterochromaffin cells (especially at the level of the lamina propria) and these cells have a high concentration of serotonin. After its secretion, serotonin metabolism is mainly hepatic, although it is also renal and pulmonary [13]. 5-Hydroxyindoleacetic Acid (5-HIAA) is the main metabolite of serotonin. It is mainly secreted in the urine, and its urinary levels are in direct relation to body serotonin levels. Urinary 5-HIAA levels are stable during the day and are not influenced by gender.

Due to the focal inflammatory infiltrate of the lamina propria characteristic of AA, it has been postulated that the release of serotonin may be increased in AA and thus its excretion at the renal level. Previous animal models confirmed that in rabbits an AA provocation model resulted in significantly higher levels of 5-HIAA than in controls [14]. This review aims to synthesize the existing evidence regarding the diagnostic ability of 5-HIAA in urine in AA.

## Methods

### Literature search and selection

We followed the Preferred Reporting Items for Systematic Reviews and Meta-Analyses (PRISMA) guidance. We specifically designed and implemented a review protocol that was registered in the international prospective register of systematic reviews (PROSPERO ID CRD42023399541). Eligible studies were identified by searching the main existing medical bibliography databases (PubMed, Medline, OVID, Web of Science, Scopus, and Cochrane Library). Search terms used for medical subject headings and keywords were: (appendicitis OR acute appendicitis) AND (5-HIAA OR 5-Hydroxyindoleacetic acid OR serotonin metabolite). The search was last executed on 13.03.2023.

Inclusion and exclusion criteria are shown in Supplementary File 1. JAM and OBA made the selection of articles. Disagreements were resolved by consensus.

### Quality assessment

The selected articles ’ methodological quality and risk of bias were evaluated with the QUADAS2 tool. Patient selection, index test, reference standard, and flow and timing were evaluated in each selected article. Applicability concerns regarding patient selection, index tests, and reference standards were also assessed.

### Data extraction and synthesis

Three independent reviewers (JAM, OBA, and BPR) extracted the relevant data from the selected articles following a standardized procedure. Extracted data included author, year of publication, study design, study population (sample size, age range, and sex distribution), AA group and control group definitions, mean and standard deviation (or median and interquartile range) for urinary 5-HIAA, statistical p-value for the between-group comparison, true positives rate, false positives rate, true negatives rate, false negatives rate, 5-HIAA cut-off value (if established), and its associated sensitivity and specificity. There were no disagreements between the reviewers after collating the extracted data. The metrics used in each study were reviewed and a standardization of units was performed when necessary. Conversion from mg/L to μmol/L was performed when necessary following molar mass references from Sigma Aldrich, Human Metabolome Database, and ChemSpider for 5-HIAA (191.183 g/mol).

### Meta-analysis

A random-effects meta-analysis for urinary 5-HIAA (AA vs control group) was performed. The results were plotted in a forest plot. Between-study heterogeneity was assessed using the Chi^2^ and I^2^ statistics. Additionally, a diagnostic test accuracy meta-analysis (DTA) was performed. The results were plotted using an hSROC diagram.

## Results

The search returned 135 articles (Scopus n=60; Pubmed n=36; Web of Science n=29; OVID n=10). Forty-five duplicates were removed. Among the remaining 90 articles, we excluded 78 following the inclusion and exclusion criteria. This review finally included 12 studies with data from 1467 participants (724 patients with a confirmed diagnosis of AA and 743 controls) [15–26]. The flowchart of the search and selection process is shown in Figure 1.

**Figure 1.**
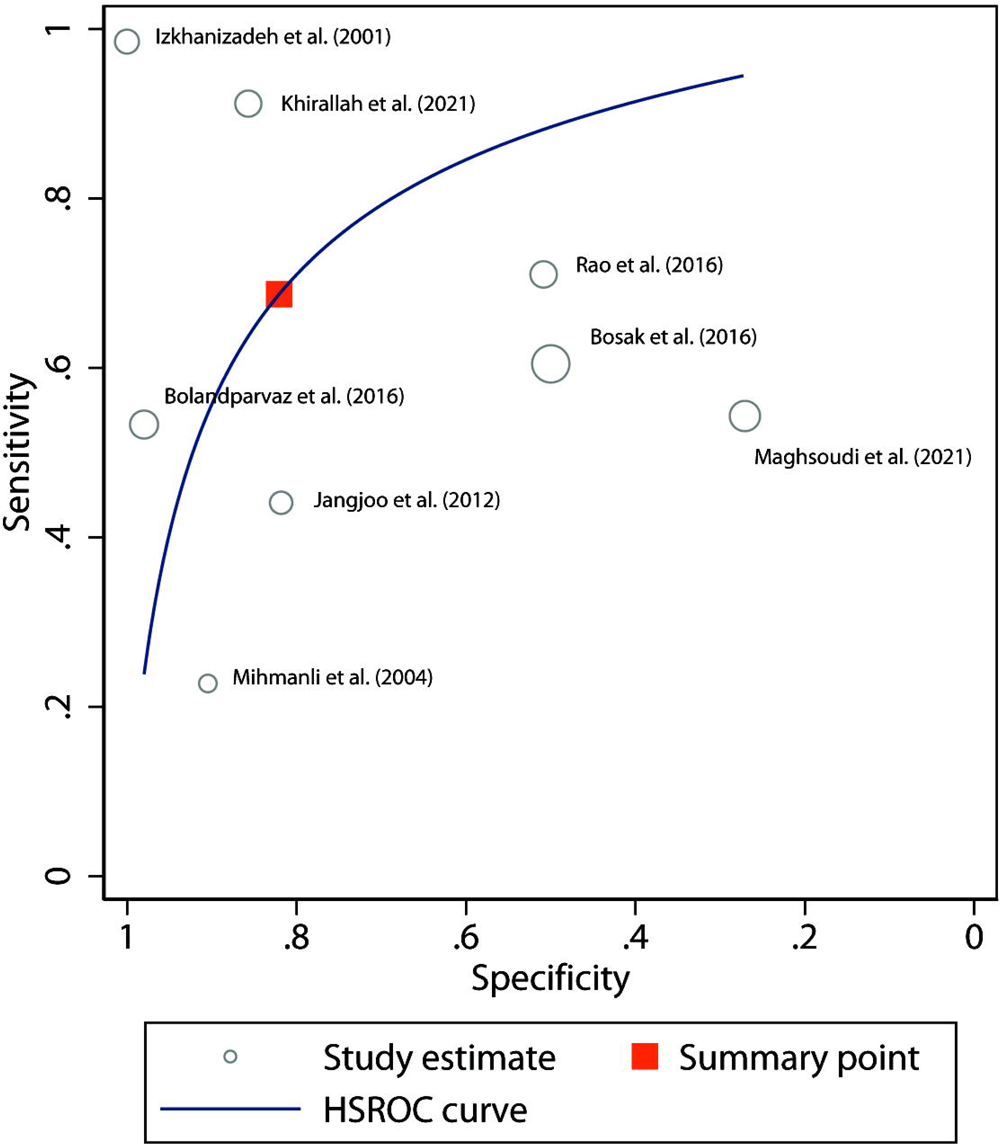
Flowchart of the search and selection process.

The risk of bias concerning the selection of patients was considered low in 6 of the 12 studies [17,18,20,21,23,25], unclear in 5 of them [15,16,19,22,26], and high in the last one [24]. The risk of bias concerning the index test was considered low in 9 of the studies [15,17,19,20–23,25,26] and unclear in 3 of them [16,18,24]. The risk of bias concerning the reference standard was considered low in 11 of the studies [15–23,25,26] and high in one of them [24]. The risk of bias concerning flow and timing was considered unclear in all the studies [15–26]. Regarding applicability concerns, the risk was estimated as low in all articles except for Kamal et al., which was found to be unclear in all three categories [24]. The results of the QUADAS2 analysis are shown in Figure 2.

**Figure 2.**
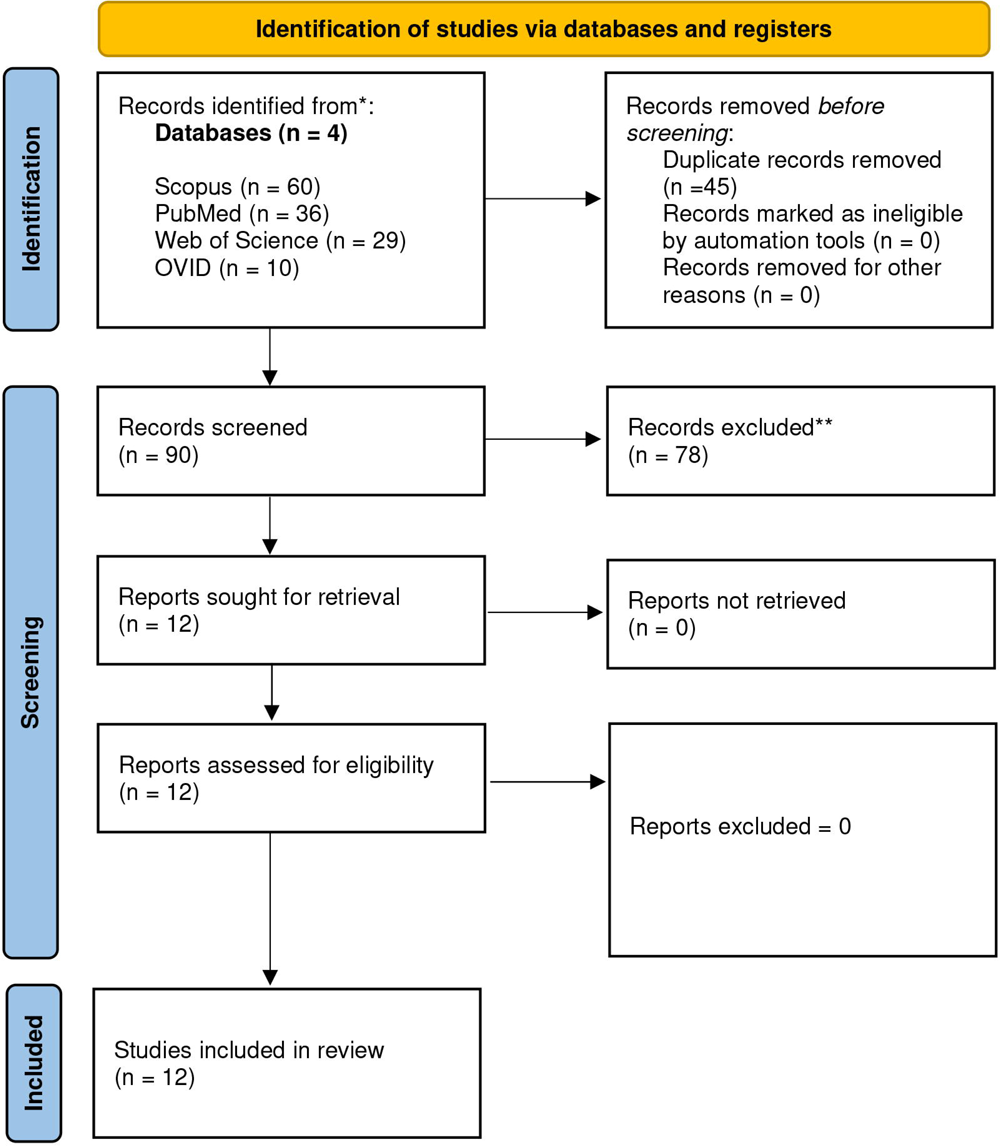
Graphical representation of the quality assessment of the diagnostic accuracy studies included in the review (QUADAS2)

### Urinary 5-Hydroxyindoleacetic Acid in acute appendicitis

#### Sociodemographic characteristics

The data extracted from the 12 studies that compared urinary 5-HIAA levels are summarized in Table 1 [15–26]. All studies were carried out between 2001 and 2021. One was from Egypt [25], 1 from the United States [20], 1 from Croatia [22], 1 from the United Kingdom [23], 1 from Irak [24], 3 from Turkey [17–19] and 3 from Iran [15,16,26]. Although not explicitly stated in all of them, all studies seemed to be prospective and 2 of them were specifically reported as Randomized Control Trials [20,25]. Three of the studies involved only pediatric populations aged between 1 and 18 years [19,22,25]. In all studies, cases were recruited in the Emergency Department before diagnosis, and biological samples were obtained at the time of inclusion in the study.

“The definition of “case” was consistent in all the selected studies, given as the histopathological confirmation of AA in the surgical specimen [15–26]. This was not the case for the definition of “control”, which was constituted either non-surgical abdominal pain [15,16,17,19,20,23,25], negative appendectomies [15,16,17,19–23,25,26], healthy controls [15,16,18,19,21,22,24] and patients with other intraoperative surgical diagnoses (such as cholecystitis, ovarian cyst or transmural ileal necrosis) [16,20,26].

Ten studies specifically evaluated potential conditions that could have altered urinary 5-HIAA levels [15,16,18–23, 25–26]. Of these, 9 asked about specific foods that might have elevated 5-HIAA levels (such as avocado, banana, eggplant, tomato, and walnuts) [15,16,18–22,25,26], 7 asked about specific medications that could have elevated 5-HIAA (such as monoamine oxidase inhibitors, serotonin and norepinephrine reuptake inhibitors, lithium or aminosalicylates) [15, 18,19,20,23,25,26] and 3 asked about specific medical conditions that could have elevated 5-HIAA levels (such as gastroenteritis, cystic fibrosis, Huntington disease, Duchenne muscular dystrophy, sickle cell disease, inflammatory bowel disease, carcinoid tumors) [18,20,26] Two studies did not report this specific evaluation [17,24]

#### 5-HIAA measurement units

Seven studies reported the values as a mean (standard deviation) [15,16,17,19,21,24,25], 1 study as a median (range) [23], and 2 as a mean (95% confidence interval) [20,22]. One study did not provide a dispersion measure [26]. One study did not provide quantitative values for 5-HIAA [18]. The reported measurement units for 5-HIAA were μmol/L [15,16,20,22,23,26], μmol/dL [19], mg/L [17,21,24], and mg/g Cr [25]. In the case of Ozel et al. [19], the unit conversion we performed resulted in values 10 times higher than those presented in the other papers. We believe that this is due to an error in the units reported by the authors, and we believe that it is most likely that the values reported were μmol/L and not μmol/dL. We tried to contact the authors to resolve this question but did not obtain a response.

Following a standardized procedure, we converted the median (range) data from Rao et al. [23] to mean (standard deviation) [27]

#### Determination of urinary 5-HIAA

Five studies determined 5-HIAA using High-performance liquid chromatography (HPLC) [15,16,19,20,22] and 5 studies through an Enzyme-Linked ImmunoSorbent Assay (ELISA) [21,23–26]. Two authors reported using spectrophotometric methods [17,18].

Regarding the authors that used HPLC, 2 used the C18 Novapack Waters associates (HPLC reagents MERK company, Germany ©) [15,16], 1 used the Hewlett Packard ® 11000 series HPLC system / Clinrep ® (Recipe chemicals and instruments GmbH Munich, Germany) [19], 1 used the Bio-Rad ® Urinary 5-HIAA test kit (Hercules, Calif) [20] and 1 used the Shimadzu HPLC machine series 10 (Shimadzu Corporation ©, Kyoto, Japan) with a CLC 100 electrochemical detector (Chromsystems Instruments & Chemicals, GmbH ® Munich Germany) [22]. Regarding the authors that used ELISA, Janjoo et al. [21] reported using the 5-HIAA ELISA kit from IBL International GmbH ® Germany, Rao et al. [23] reported using the 5-HIAA ELISA kit from ALPCO ® and Khirallah et al. [25] reported using the 5-HIAA ELISA kit from LDN, DN Labor diagnostika Nord GmbH & Co ®. Nordhorn, Germany

#### Diagnostic performance of urinary 5-HIAA

Nine studies provided a p-value for the comparison between 5-HIAA values in the AA group and in the control group, 6 of them being statistically significant [15,16,19,23,24,25]. The area under the curve (AUC) reported ranged from 0.55 [22] to 0.985 [15], and the proposed cut-offs ranged from 7.4 μmol/L [26] to 27.20 μmol/L [21]. The reported sensitivity ranged from 22% [17] to 98% [15] and the reported specificity ranged from 27.1% [26] to 100% [15,24]. Although none of the work evaluated in a specific way the discriminatory capacity of this molecule between non-complicated AA (NCAA) and complicated AA (CAA), the majority of works did not find significant differences in the levels of 5-HIAA between both groups (even some groups, such as Kamal et al., found higher 5-HIAA levels in the NCAA group).

#### Random-effects meta-analysis for urinary 5-HIAA

Due to the inconsistencies found in terms of units in the work of Ozel et al. [19], we decided to exclude this work from the random effects meta-analysis. Hernandez et al. [20] and Bosak et al. [22] provided the dispersion measure of 5-HIAA as a 95% confidence interval and Maghsoudi et al. [26] provided a single 5-HIAA central tendency measurement without a dispersion measure. Lastly, Kirallah et al. [25] provided the data adjusted to urinary creatine. Therefore, we could not include this works in the meta-analysis.

The random-effect meta-analysis of urinary 5-HIAA (AA vs controls) included 7 articles (352 AA and 258 controls) and resulted in a significant mean difference [95% CI] of 23.30 [15.82-30.77] μmol/L (p<0.001). The I^2^ value was 97% and the Chi^2^ value was 234. The graphical depiction of the analysis is shown in Figure 3.

**Figure 3.**
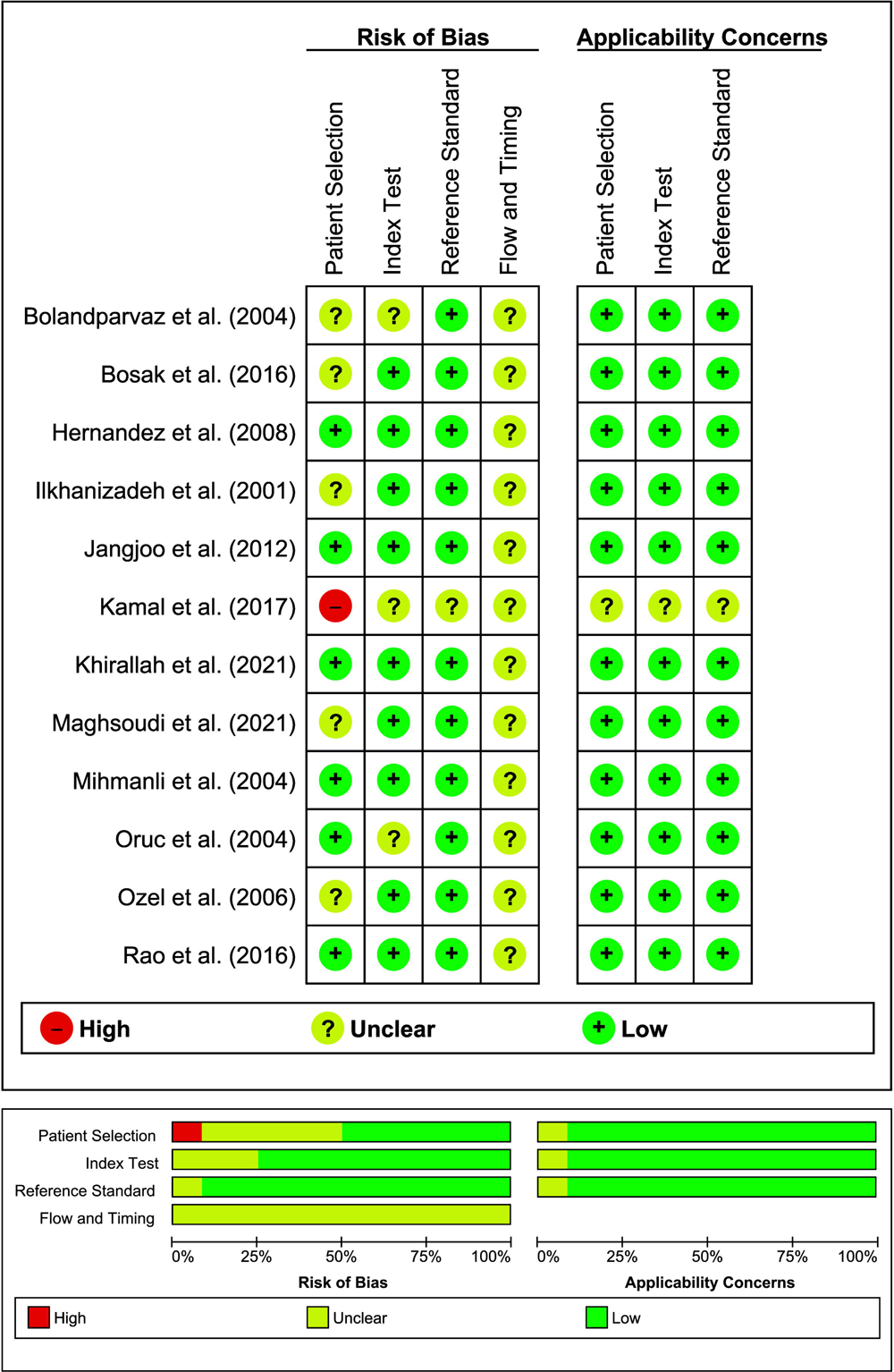
Forest plot of the random-effects meta-analysis performed for urinary 5-HIAA (AA group vs. Control group).

#### Diagnostic test accuracy meta-analysis for urinary 5-HIAA

Four authors provided true positive (TP), false positive (FP), true negative (TN), and false negative (FN) data from their studies [15,16,21,26].

We calculated TP, FP, TN, and FN data in 4 of the works with the provided data (sensitivity, specificity, predictive values) [17,22,23,25]. Although minor inconsistencies were identified in terms of decimals, probably related to numerical rounding, we believe that the data are correct.

We tried to contact through the corresponding author address and ResearchGate ® with Oruc et al. [18], Ozel et al. [19], Hernandez et al. [20], and Kamal et al. [24] to ask for TP, FP, TN, and FN data because in these cases this information was not inferable through the articles. We obtained no response.

The DTA meta-analysis of urinary 5-HIAA included 8 articles and resulted in a pooled sensitivity [95% CI] of 68.6 [44.1-85.9]% and a pooled specificity [95% CI] of 82 [54.7-94.5]%. The graphical depiction of the DTA meta-analysis is shown in Figure 4.

**Figure 4.**
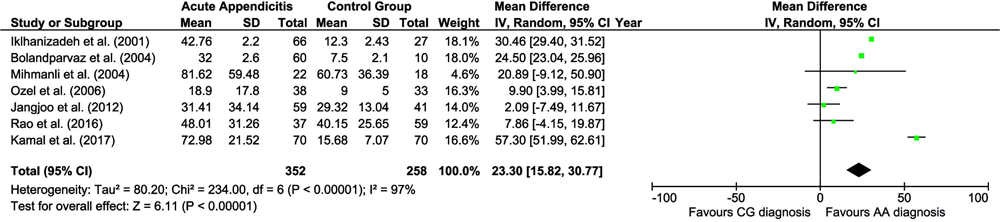
hsROC plot of the DTA meta-analysis performed for urinary 5-HIAA (AA group vs. Control group).

## Discussion

This study systematically reviewed the evidence on the role of urinary 5-HIAA in the diagnosis of AA. We synthesized the results of 12 prospective studies including 724 patients with a confirmed diagnosis of AA and 743 controls, and performed 2 different meta-analyses: First, a random-effects meta-analysis that showed significantly higher mean values of urinary 5-HIAA in the AA group than in the control group; second, a DTA meta-analysis that showed a low-to-moderate pooled sensitivity (68.6%) and a relatively high pooled specificity (82%).

These findings, even with a limited degree of evidence, suggest that urinary 5-HIAA could be a promising marker for the diagnosis of AA.

Regarding the diagnostic performance of 5-HIAA for the discrimination between AA and controls, the wide range of AUC, sensitivity, and specificity reported are striking. A reasonable justification for this variability is the heterogeneity in the control groups of the different studies (including healthy patients, negative appendectomies, other intraoperative surgical diagnoses, and non-surgical abdominal pain). The presence of a control group of healthy patients will always tend to overstate the diagnostic performance of the biomarker in this context, compared to a control group of patients with acute abdominal pain, in whom there may always be a certain level of systemic inflammation and metabolic stress that leads to an elevation of the biomarker. This heterogeneity in the control groups would also explain the high between-study heterogeneity observed in the meta-analysis (I^2^=97%).

We believe that the pathophysiological premise that justifies the evaluation of 5-HIAA as a potential diagnostic marker in AA is valid, as demonstrated by the random effects meta-analysis performed: urinary 5-HIAA levels are significantly higher in the AA group than in the control group in the meta-analytical model proposed. However, this does not necessarily translate into adequate diagnostic performance. Because of this, we performed a specific DTA-type meta-analysis. It should be noted that there is a potential selection bias in this analysis, given that 4 of the 12 papers in this review were not included due to a lack of data.

When we explored the potential sources of heterogeneity in this work, we found (apart from the selection of the control group) an important variability in the determination techniques employed: HPLC and ELISA are radically different techniques both in terms of precision and cost. We believe that whenever a diagnostic test is evaluated in the context of a prevalent and time-dependent pathology, such as AA, a simple, inexpensive, and rapid determination methodology should be primed, and in this case, ELISA is probably the most appropriate technique. On the other hand, it is curious that except for the work of Bosak et al. [22], which did use HPLC, since 2012 all the reported works have used ELISA. However, we have not found any chronological pattern in the diagnostic yields published for this molecule in this review.

Another aspect to consider when evaluating a diagnostic test is the limitations that the patient’s baseline situation may present for its use. For example, Ca 125 has been evaluated as a potential biomarker in the context of AA, but its potential elevation due to multiple other causes (such as gynecological pathology) justifies its poor diagnostic performance [28]. In this case, it should be considered that multiple medical, pharmacological, and dietary conditions can elevate urinary 5-HIAA levels, and this is a major limitation to the use of this molecule as a routine diagnostic test in AA.

During the course of this work, we have found inconsistencies in various papers that should be explicitly mentioned to adequately interpret the results. In the case of Jangjoo et al. [23], the authors report having excluded from the analysis the outliers’ values of 5-HIAA obtained. This, in our opinion, is an important source of bias, given that many previously reported biomarkers, such as serum IL-6, may present extreme values in the context of AA and should be known for a proper understanding of the behavior of the molecule in this disease [4]. Additionally, it is pertinent to highlight that some of the studies do not adequately clarify what happens at the classification level with negative appendectomies or with patients who are initially classified as non-surgical abdominal pain and subsequently develop acute appendicitis, which constitutes an important source of confusion [25].

The main strengths of this work are its rigorous methodology, following international PRISMA guidelines and applying the QUADAS2 bias assessment tool [29,30], and the use of a specific meta-analytical model, such as the DTA. However, this study is not exempt from limitations. 1) the sample size included is limited and the populations are heterogeneous. 2) not all the working groups have considered the potential factors that could have elevated urinary 5-HIAA 3) The absence of data has led to a potential bias when performing the meta-analyses since not all the studies have been included 4) Although in the case of urinary LRG1, multiple studies reported their results adjusted for urinary creatinine, in this review only 1 publication made this adjustment 5) a small percentage of the numerical data included in this study have been obtained using an inferential methodology and there may some inaccuracies.

In conclusion, although the evidence is very limited, urinary 5-HIAA is emerging as a potential diagnostic tool for AA. Future prospective studies with a large sample size and with an adequate assessment of factors that may elevate serotonin before the determination of urinary 5-HIAA levels should be conducted. Urinary 5-HIAA adjustment for creatinine, previously reported in the case of urinary LRG1, is an interesting aspect to evaluate in the future for this type of diagnostic biomarker. Urinary 5-HIAA does not seem to be a useful biomarker to distinguish between NCAA and CAA.

### CRediT authorship contribution statement

**JAM:** Conceptualization and study design; literature search and selection, data curation and extraction, formal analysis; investigation; methodology; project administration; resources; validation; visualization; writing – original draft; writing – review and editing.

**OEB, BPR, MRJ:** literature search and selection, data curation and extraction, visualization; writing – reviewing, and editing.

## CONFLICTS OF INTEREST

The authors declare that they have no conflict of interest.

## FINANCIAL STATEMENT/FUNDING

This review did not receive any specific grant from funding agencies in the public, commercial, or not-for-profit sectors. None of the authors have external funding to declare.

## ETHICAL APPROVAL

This study did not involve the participation of human or animal subjects, and therefore was exempt from formal assessment by the ethics committee for clinical research of our center.

## STATEMENT OF AVAILABILITY OF THE DATA USED DURING THE SYSTEMATIC REVIEW

The data used to carry out this systematic review are available upon request from the review authors.

## Supporting information

PRISMA checklist

## Data Availability

All the data pertaining to this systematic review is available upon reasonable request through the corresponding author

## Notes

### Competing Interest Statement

The authors have declared no competing interest.

### Funding Statement

This study did not receive any funding

