## Supplementary File 1 for "Diagnostic performance of urinary 5-Hydroxyindoleacetic Acid in acute appendicitis: a systematic review and diagnostic test accuracy meta-analysis"

**Exclusion criteria**

-Case reports.

-Duplicate or overlapping studies.

-Reviews, systematic reviews, consensus guidelines.

-Languages other than English or Spanish.

-Studies with no surgical intervention.

-Studies with no population of interest.

-Studies conducted in immunocompromised patients.

-Studies conducted in patients with metastatic neoplastic disease and invasive abdominal neoplastic disease.

-Studies conducted in patients with acute or chronic kidney disease.

Studies conducted in patients with dietary, pharmacological or medical conditions involving a potential baseline elevation of 5-Hydroxyindoleacetic Acid.

**Inclusion criteria**

-Prospective or retrospective observational original clinical studies evaluating the diagnostic accuracy of urinary 5-Hydroxyindoleacetic Acid in relation to the reference standards for the diagnosis of appendicitis and/or for the discrimination between complicated and uncomplicated appendicitis.

-Diagnostic validation studies evaluating the diagnostic accuracy of urinary 5-Hydroxyindoleacetic Acid in relation to the reference standards for the diagnosis of appendicitis and/or for the discrimination between complicated and uncomplicated appendicitis.

**Supplementary file 1. Inclusion and exclusion criteria**
